# A machine-learning evaluation of biomarkers designed for the future of precision medicine

**DOI:** 10.1101/2023.07.09.23292430

**Authors:** Sharlee Climer

## Abstract

Precision medicine is cognizant of the impact of genetics and environments on subtypes of heterogeneous diseases and aims to identify, diagnose, and treat each subtype appropriately. Real-valued biomarkers, such as protein levels in plasma, are key for practical subtype diagnoses and hold potential to elucidate subtypes and illuminate promising drug targets. Biomarkers that are common across all subtypes have been discovered using fold change (FC) and the area under the receiver operating characteristic curve (AUC). However, FC and AUC fail to identify biomarkers for subtypes when they comprise less than half of the disease group. We present here a machine-learning biomarker evaluation method based on clustering of the data points, referred to as Difference in Bicluster Distances (DBD). We contribute efficient, yet optimal, software coupled with rigorous validation techniques, and demonstrate our approach on a late-onset Alzheimer disease (AD) gene expression dataset. Our trials produced four significant genes and appropriate thresholds for biomarker diagnostics. While none of these genes were identified as significant by either FC or AUC for the given dataset, the genes have been independently associated with AD or neurological disorders by other groups using completely independent means. In summary, DBD provides a unique and effective method for screening real-valued data to identify biomarkers associated with subtypes of heterogeneous diseases.

## Introduction

Many complex diseases of interest are heterogeneous, in that they arise due to multiple distinct causes, yielding various subtypes. The goal of precision medicine is to identify, diagnose, and develop treatments for each subtype. Many successes have been made, such as the identification of distinct subtypes of breast cancer, e.g. luminal A, luminal B, HER2 positive, and basal-like, and treating each subtype appropriately. However, progress toward developing precision medicine for many diseases plaguing humanity, such as late-onset Alzheimer disease (AD), has been slow. AD is a heterogeneous disease, as revealed by its wide range of genetic and environmental risk factors and diversity of clinical manifestations, yet the lack of knowledge about these subtypes impedes development of treatments to cure this devastating disease. Drug discovery and selections of individuals for drug trials are severely handicapped by the lack of ability to discern subtypes.

Most complex diseases arise due to combinations of genetic and environmental factors, such as lifestyle. In addition to genetic testing, biomarkers generated by procedures such as PET or MRI scans, biopsies, lumbar punctures, and plasma sampling provide clues for identifying subtypes. Many of these procedures produce real-valued data, such as levels of proteins in plasma or cerebral spinal fluid, and amyloid or tau loads in various brain regions. Real-valued biomarkers are commonly evaluated using either fold change (FC) or the area under the receiver operating characteristic curve (AUC). FC is the ratio of the mean or median of biomarker levels for the diseased cases and normal controls. AUC is the area under the curve of a 2-dimensional plot of the true positive rate vs. the false positive rate as these values range from zero to one.

We previously demonstrated the inability for FC and AUC to capture signals for subtypes when they comprise less than 50% of the diseased individuals.^1^ FC is unable to capture the subtypes as only the mean or median is used in the computation and subtype signals are lost. AUC’s dependence upon the true positive rate cripples its evaluation as the upper limit on this rate is the percentage of individuals in the subset. As most heterogeneous disease subtypes are expected to represent less than half of the diseased individuals, FC and AUC are inappropriate metrics for evaluating biomarkers for precision medicine.

Cognizant of the limitations of the true positive rate, as well as the related measurements in the confusion matrix, for detecting subtypes we previously proposed a bimodality coefficient difference (BCD) evaluation.^1^ Based on the assumption that a subtype will create a secondary peak in the distribution of the data for diseased cases, BCD computes the bimodality coefficient for each group of diseased cases and normal controls and is set to the difference of these two values. The bimodality coefficient is based on statistical characteristics of the data, including skewness, cardinality, and kurtosis.^2^ Using BCD, we presented dramatic improvement over AUC and FC for large sample sizes, including thousands of simulated trials with 1000 individuals and RNA sequencing of ∼2k Mus musculus microglia cells tracked during neurodegeneration.

During the course of our trials, we have observed that statistical characteristics indicative of bimodality tend to erode with smaller sample sizes. Due to the limited numbers of individuals in many research trials due to budget constraints, we present herein an alternative approach based on the concept that data points in the two modes resemble two natural clusters.

Clustering of data points has been heavily researched in the field of machine learning. *k*-means and *k*-medians are two methods that minimize the sum of the distances of the points to the means or medians, respectively, of their assigned clusters. We opt for *k*-medians due to potential issues of using means in the presence of extreme values and/or outliers. Given a set of points, *k*-medians aims to subdivide the points into *k* subsets such that the following summation of squares is minimized:

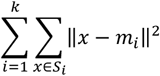

where *k* is the number of clusters, *Si* is the set of points that are in cluster *i*, and *mi* is the median for cluster *i*.

Optimally solving *k*-medians is generally NP-hard^3,4^ and Lloyd’s algorithm^5^ is commonly used to identify an approximate solution. Lloyd’s algorithm is an expectation-maximization method in which data points are first assigned to arbitrary clusters, then two alternating steps are iteratively repeated until data points no longer move to different clusters. The two steps are (1) medians are computed for each cluster and (2) the points are reassigned to their closest median. The closeness between two points can be assessed using metrics such as Euclidean distance. In general, the number of clusters is typically unknown prior to the analysis and multiple trials are run with sequential values for *k*. For our purposes, *k* is always set to two (*2*-medians) as we aim to see how well the data points clump into two clusters.

We assume that an analyte associated with a subtype for a disease will have values that tend to form two clusters for the diseased cases, in which one cluster represents the values for the individuals with the subtype. Greater separation between the medians for the two clusters represents higher distinction between the subtype and the other diseased cases. Note that it is possible that the normal controls generate two clusters for analytes that naturally vary, such as glucose levels in blood. For this reason, we compare how well the diseased cases vs. the normal controls cluster into two distinct clusters.

We present Difference in BiCluster Distances (DBD) for evaluating real-valued biomarkers for heterogeneous diseases. First, the samples are split into Discovery and Validation datasets. Candidate biomarkers are identified by analyzing the Discovery data and then tested in the unseen Validation data to ensure reliability of the results. Second, each analyte is evaluated by computing the optimal *2*-medians solution for the diseased cases and then for the normal controls in the Discovery dataset. The distance between the medians for the controls is subtracted from the distance between the medians for the cases and this difference is recorded as *DBD*_*i*_ for analyte *i*. Third, each *DBD*_*i*_ is evaluated for significance by running a series of permutation trials in which the values for the analyte are randomly assigned to a case or control label and the *DBD* value is computed. This procedure breaks down any associations that might exist and provides a p-value for each analyte. Fourth, the most significant analytes are tested on the individuals in the Validation dataset and corrections for multiple testing are applied. We conservatively utilize Bonferroni corrections to provide high confidence in final results.

Optimally solving *k*-medians is NP-hard^3,4^, but we were able to develop a highly efficient optimal algorithm with worst-case time complexity of *O*(*mn*^2^), where *m* is the number of analytes and *n* is the number of individuals in the group with highest cardinality. This efficiency is possible due to the low dimensionality of the data points, setting *k* to 2, and using a unique dynamic programming approach which stores subproblem solutions for reuse.

In addition to introducing the DBD method, we present results from using this method on a gene expression dataset comprised of 7,431 genes for 173 AD cases and 184 normal controls. These trials yielded four significant genes after Bonferroni corrections when tested on the independent Discovery data. None of the four genes are significant when analyzed using AUC or FC. DBD provided thresholds for direct translation to biomarker trials and these thresholds exhibit extremely strong odds ratios.

## Methods

We utilized previously generated Sentrix HumanRef-8 Expression BeadChip22 gene expression data (GEO Omnibus GSE15222), which consists of expression levels for 8,560 genes derived from human cortex tissue of 176 AD cases and 188 age-matched controls.^6^ The dataset was cleaned to a maximum of 5% missing values per individual and per gene using the DataRetainer program (http://www.cs.umsl.edu/~climer/blocBuster/code.html). The cleaned data are comprised of 7,431 genes for 173 AD cases and 184 controls and are available by request.

The overall procedure is tabulated in **Figure 1**. Optimal *2*-medians clustering was implemented using a dynamic programming algorithm, which eliminates redundant computations while ensuring the optimal solution is returned. Quicksort was utilized for sorting data. Ten thousand permutation trials were performed for each analyte, providing accurate p-values with four significant digits. This large number of permutation trials also ensures adequate sample size for analytes returned as significant.

**Figure 1.**
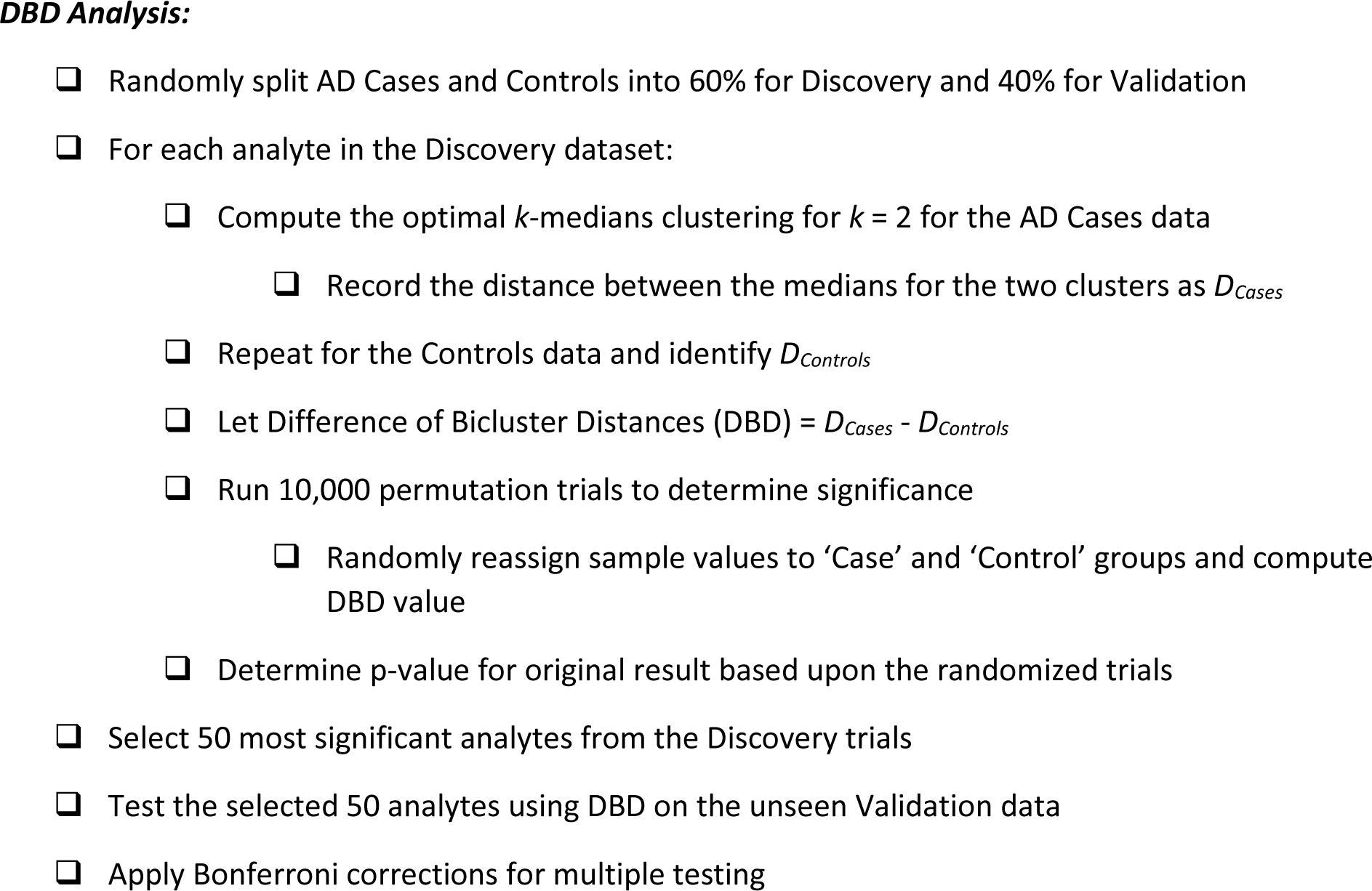
Steps used in our data analysis of gene expression data for AD cases and normal controls.

## Results

When tested on the unseen Validation data, 15 of the 50 genes with the highest significance in the Discovery trials had p-values ≤ 0.05. Four of these 15 genes had p-values ≤ 0.05 after Bonferroni corrections for multiple testing (**Table 1**).

**Table 1.** DBD, AUC, FC, and gene information for the four genes identified by DBD. FC shown is the absolute value of the log2 of the ratio of case/controls medians.

| Gene<br>Symbol | DBD |  |  | AUC | FC | Description | Refseq | Cytoband |
| --- | --- | --- | --- | --- | --- | --- | --- | --- |
|  | Discovery<br>p-value | Validation<br>p-value | Bonferroni<br>corrected |  |  |  |  |  |
| *UBE2H* | 0.0002 | 0.0002 | 0.010 | 0.546 | 0.042 | ubiquitin-conjugating<br>enzyme E2H (UBC8<br>homolog, yeast) | NM_182697.1 | 7q32 |
| *FEZ1* | 0.0008 | 0.0005 | 0.025 | 0.585 | 0.187 | fasciculation and<br>elongation protein<br>zeta 1 (zygin I) | NM_022549.2 | 11q24.2 |
| *TMEM5* | 0.0004 | 0.0007 | 0.035 | 0.540 | 0.026 | transmembrane<br>protein 5 | NM_014254.1 | 12q14.2 |
| *APBA2* | 0.0005 | 0.0010 | 0.050 | 0.566 | 0.145 | amyloid beta (A4)<br>precursor protein-binding, family A,<br>member 2 | NM_005503.2 | 15q11-q12 |

The four significant genes are: ubiquitin-conjugating enzyme E2H (*UBE2H*), fasciculation and elongation protein zeta 1 (*FEZ1*), transmembrane protein 5 (*TMEM5*), and amyloid beta precursor protein-binding family A, member 2 (*APBA2*). Histograms of these four genes illuminate subtypes within the AD Cases (**Figure 2**).

**Figure 2.**
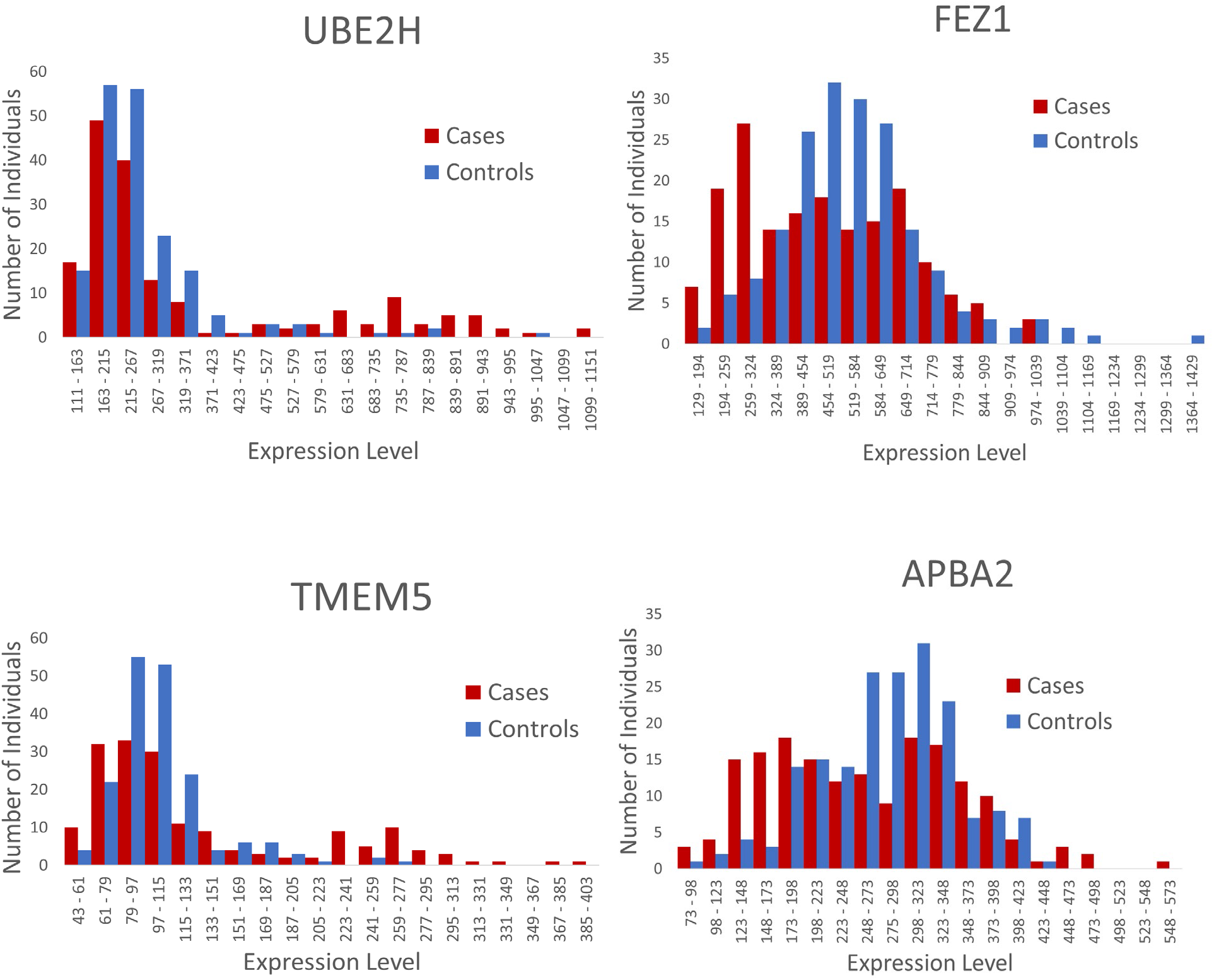
Histograms for the four genes identified by DBD.

Note that none of the four genes are significant when analyzed using AUC or FC (**Table 1**). Thresholds suggested by the DBD results yield extremely strong odds ratios, as shown in **Table 2**. DBD directly provides thresholds that can be utilized in biomarker diagnostics.

**Table 2.** Thresholds suggested by the DBD results, percentages of AD cases and normal controls with corresponding values, odds ratio, and 95% confidence interval for the odds ratio.

| **Gene Symbol** | **Threshold** | **% Cases** | **% Controls** | **OR [95% CI]** |
| --- | --- | --- | --- | --- |
| *UBE2H* | >=600 | 22.5% | 2.7% | 10.4 [4.0, 27.1] |
| *FEZ1* | <= 325 | 30.6% | 8.7% | 4.6 [2.5, 8.5] |
| *TMEM5* | >=220 | 21.1% | 1.7% | 15.8 [4.8, 52.5] |
| *APBA2* | < =174 | 23.7% | 6.0% | 4.9 [2.4, 9.9] |

Total computation time for the Discovery trials was 24 minutes. Over 74 million DBD computations were conducted, with an average time of 1.9 × 10^−5^ seconds per evaluation. This efficiency is due to the use of a unique dynamic programming algorithm, fast sorting algorithm, and C/C++ code implementation.

## Conclusions

DBD provides a machine-learning approach for assessing real-valued biomarkers. It has no reliance upon statistical characteristics of the data making it robust for practical sample sizes. Also, it utilizes medians, rather than means, thereby providing robustness in the presence of outliers and other extreme values.

Although optimally solving *k*-medians is NP-hard, our *2*-medians objective over 1-dimensional data using dynamic programming is highly efficient and enables the use of a large number of permutation trials to determine the significance for each analyte. The permutation trials retain the original data values and provide insights into the likelihood of observing a given DBD value for the specific data values.

We demonstrate the utility of our approach by revealing four genes worthy of further investigation. It is important to note that both AUC and FC failed to identify any of these four genes for the given dataset, yet independent research by others reveal associations. *UBE2H* has previously been identified as a potential biomarker for AD.^7^ Also, it was recently shown that reversal of increased FEZ1 in rats with induced AD suggests a mechanism for the effects of melatonin.^8^ Although a brief search did not reveal any direct associations between AD and TMEM5, transmembrane proteins in general play many roles in neurological disorders.^9^ Finally, *APBA2* encodes a protein that interacts with the Alzheimer’s disease amyloid precursor protein, APP, which is cleaved during the production of amyloid beta, and several studies suggest that APBA2 regulates amyloid beta production.^10–13^

It is patently clear that AUC and FC fail to identify biomarkers for subtypes of heterogeneous diseases when the subtype comprises less than half of the entire group of diseased cases.^1^ Subtypes have inherently low true positive rates, which sabotage AUC assessments, and are lost in summary statistics such as FC. Instead of focusing on these traditional measurements, we have developed tools based on the assumption that a subtype will form a secondary cluster within the data values for the diseased cases. Our recent approach, BCD, is based upon the statistical characteristics of the data and shows great improvements for large sample sizes.^1^ This manuscript introduces a machine-learning approach, DBD, based on clustering that is suitable for more moderate sample sizes. In summary, DBD provides a unique and effective method for screening real-valued data to identify biomarkers associated with subtypes of heterogeneous diseases.

## Data Availability

https://www.ncbi.nlm.nih.gov/geo/query/acc.cgi?acc=GSE15222

## Acknowledgements

This research was funded by the Alzheimer’s Association (AARG-22-925002) and research grants from the University of Missouri – St. Louis.

